# Regulatory Myeloid-Cell Dynamics In Asthmatics During Pregnancy

**DOI:** 10.1101/2025.05.23.25328090

**Authors:** Sultan Tousif, Yong Wang, Zhen Chen, Tzu-Chun Lin, Shaheer Ahmad, Seth Sherman, Mickey Parks, Jennifer Trevor, Jennifer Weck, Joseph R. Biggio, Akila Subramaniam, Pauline Mendola, Jessy S. Deshane

**Affiliations:** Department of Medicine, University of Alabama at Birmingham, Birmingham, AL; Division of Intramural Population Health Research, Eunice Kennedy Shriver National Institute of Child Health and Human Development, Bethesda MD; The Emmes Company, LLC, Rockville, MD; Department of Obstetrics and Gynecology, University of Alabama at Birmingham.; Department of Epidemiology and Environmental Health, School of Public Health and Health Professions, University at Buffalo, Buffalo NY; Department of Cellular Biology & Anatomy, LSUHS | LSU Health Shreveport

## Abstract

Asthma is one of the most common chronic inflammatory lung diseases among pregnant women. Acute asthma exacerbations during pregnancy negatively impact pregnancy outcomes. As myeloid-derived suppressor cell (MDSC) populations are known regulators of airway inflammation and are important in maternal-fetal tolerance, we investigated if percentages of circulating MDSC subsets correlated with clinical indicators of airway inflammation and maternal-fetal tolerance in a cohort of healthy pregnant (HP), asthmatic pregnant (AP) and well controlled asthmatic pregnant (WCAP) women recruited to the B-WELL Mom study. Reciprocal dynamics of circulating MDSC subsets, with granulocytic MDSCs (Gr-MDSCs) increasing over early pregnancy and declining in late stage pregnancy and the reverse trend with monocytic MDSCs (Mo-MDSCs) were identified; the decline of Mo-MDSCs at Visit 3 of pregnancy was higher in AP/WCAP compared to HP whereas the decline of Gr-MDSCs at late stage pregnancy was lower in AP/WCAP compared to HP. In AP/WCAP, positive correlations were observed between both MDSC subsets during pregnancy and Gr-MDSCs correlated positively with fractional exhaled nitric oxide, a surrogate measure of airway inflammation, during early pregnancy; these relations were not identified in HP subjects. Interestingly, soluble HLA-G as well as indoleamine 2,3 dioxygenase activity, both important in maternal-fetal tolerance, showed positive correlations with MDSCs only in HP and not in AP/WCAP subjects. Our studies suggest that differential myeloid-cell dynamics may serve as biomarkers of maternal-fetal tolerance in AP/WCAP subjects.

## Introduction

Asthma is a chronic inflammatory disease characterized by airway inflammation and epithelial cell-crosstalk, with both innate and adaptive immune cells contributing to the reversible airway obstruction associated with airway hyperresponsiveness (AHR) (1–3). The airway infiltration of Th2 cells and innate lymphoid type 2 cells are main drivers of eosinophilic asthma (4–11). Non-eosinophilic asthma is predominantly neutrophilic, and involves the Th17 subset of helper T cells (2,12–15). In addition to Th and innate lymphoid cells, heterogeneous regulatory myeloid lineage cells have gained recognition in their role in regulating allergic airway inflammation (16–24). Subsets of myeloid-derived suppressor cells (MDSCs) with T cell suppressive potential have been recognized in many pathologic conditions (16). In humans, circulating MDSCs consist of monocytic MDSCs (Mo-MDSCs) and granulocytic MDSCs (Gr-MDSCs) (25). MDSC-like cells were found to accumulate in allergic asthma and to suppress Th2 response in TLR4 and MyD88-dependent manner, mediated by interleukin −10 (IL-10) and arginase 1 (ARG1) (24). MDSCs and IL-10 levels significantly increased and negatively correlated with IL-12 levels during the onset of asthma in both human and mice (26,27). We have previously reported the infiltration of Gr-1^+^CD11b^+^ myeloid cell subsets in murine lungs of mice during allergic airway inflammation where they differentially generated the reactive free radicals nitric oxide (NO) and superoxide (O_2_^• –^)(22,23). The monocytic subset (predominant NO producer) and the granulocytic subset suppressed T-cell proliferation *in vitro*. In contrast, the O_2_^• −^-generating macrophage-like subset have been shown to enhance T-cell responses (22), thus defining a regulatory role for MDSCs in airway inflammation. We have also identified similar pro-inflammatory superoxide-producing cells in the airways of humans that stimulated autologous T cells, and NO producing monocyte like sub populations of cells that suppressed proliferation autologous peripheral T cells. Although the pro-inflammatory subset of cells was identified in the airways of patients with mild asthma or COPD and not in circulation, their percentages discriminated these study subjects from those of healthy controls (17).

Asthma is one of the most common chronic medical conditions reported during pregnancy (28–33). In the United States, 5-8% of pregnant women and 13% worldwide, experience changes in asthma control (34–36). Asthma phenotype also influences the course of asthma during pregnancy, with higher exacerbation frequency reported in women with non-eosinophilic asthma. Up to 20% of pregnant women with asthma have at least one asthma exacerbation during pregnancy and one-third of them become hospitalized [5]. Maternal asthma is associated with an increased risk of adverse perinatal outcomes (33–39). Understanding the underlying mechanisms that contribute to these changes in asthma control is essential for preventing adverse pregnancy outcomes, and risk of newborn mortality.

Maternal–fetal tolerance is important for normal pregnancy; immune cell dynamics and functions alter dynamically in the uterus throughout pregnancy. During pregnancy, the maternal immune system tolerates the semiallogeneic fetus expressing paternal antigens without immune rejection. This requires changes/adaption in the maternal immune response, particularly the induction of limited immunosuppression without loss of antibacterial host defense (40–42). Dysfunction of the immune system can also occur during pregnancy and can lead to loss of pregnancy or severe complications (43,44). Thus, maternal-fetal tolerance has to be tightly regulated, but the underlying mechanisms are not well understood. Besides macrophages, uterine natural killer cells and different T cell subsets, myeloid-derived suppressor cells (MDSCs) have emerged as important players in fetal immune tolerance and maintenance of pregnancy. Expansion of MDSC populations were observed in peripheral blood, placenta and uterus during pregnancy in mice and in healthy women. In healthy human pregnancies, an accumulation of Gr-MDSCs as well as Mo-MDSCs have been reported with up to tenfold higher numbers of Gr-MDSC in the peripheral blood of pregnant women compared to healthy non-pregnant controls (45,46). Frequencies of Gr-MDSCs were highest during early gestation (45,46), and dropped within a few days postpartum to levels of non-pregnant women (45,46). In human placenta Gr-MDSCs were shown to be enriched in comparison to maternal and fetal blood. The maternal origin of these cells have been revealed by genetic analyses (45–48).

MDSCs exert immune regulation through multiple mechanisms, including inducible nitric oxide synthase (iNOS), ARG1, reactive oxygen species (ROS), cyclooxygenase-2 (COX-2), IL-10, TGF-β and indoleamine 2,3-dioxygenase (IDO) (25). In pregnant women, MDSCs suppress T cells via ARG1, iNOS, ROS and IDO, which promotes maternal–fetal tolerance (25,49). IDO is a heme-containing enzyme that catalyzes the conversion of the essential amino acid tryptophan (Trp) into the metabolic byproduct kynurenine (Kyn), expression of which at the maternal-fetal interface is necessary to prevent immunological rejection of fetal allografts (50–52). The IDO pathway has been found to contribute substantially to the control of asthma (53). Low IDO activity has been observed in asthmatics (54–56). The induction of IDO by certain TLR ligands inhibits Th2-driven experimental asthma (56). However, it remains to be determined how IDO activity changes in asthma during pregnancy. But the potential role of MDSCs in regulation of asthma during pregnancy remains largely unknown.

Human leucocyte antigen G (HLA-G) is a major histocompatibility complex (MHC) I molecule with immuno-modulatory properties. HLA-G is expressed during pregnancy, which plays an important role in mediating maternal-fetal tolerance (57,58). Soluble HLA-G (sHLA-G) is increased in maternal serum and reduced in patients with preeclampsia and miscarriages (59,60). It has been demonstrated that sHLA-G plays a role in Gr-MDSCs accumulation during pregnancy (48). Further, it has been shown that sHLA-G induces an upregulation of IDO expression and phosphorylation of STAT3 in myeloid cells (48). However, it is not yet known how sHLA-G changes in asthma during pregnancy.

In this study, we have analyzed percentages of circulating MDSC subsets from 64 study subjects with asthma (including asthmatic pregnant and well controlled asthmatic pregnant (AP/WCAP) and 25 healthy controls from three clinic visits during pregnancy (0 days of gestation, 20-22 weeks of gestation, 30-32 weeks of gestation) and one at 4 months post-partum. Our studies indicate correlation signatures of percentages of circulating Gr-MDSCs, Mo-MDSCs and levels of sHLA-G, IDO activity and FeNO are different between AP/WCAP and HP subjects. A further understanding of MDSC subpopulations in the pathogenesis of asthma during pregnancy might lead to identification of new biomarkers or therapeutic targets in asthmatic pregnant subjects.

## Materials and Methods

### Study Design and Subjects

B-WELL-Mom is an observational cohort designed to evaluate biologic mechanisms (allergy, genetic, immunologic) underlying changes in asthma control during pregnancy. To be eligible, women were required to be aged 18 years or older, were less than 15 weeks gestation, English or Spanish speaking, had to have a singleton pregnancy, not planning to terminate their pregnancy, planning to deliver at the study hospital, be willing to have blood drawn at each visit and without autoimmune disease (HIV, Lupus, Rheumatoid Arthritis, Multiple Sclerosis, Mixed Connective Tissue Disease) or Cystic Fibrosis. At the University of Alabama at Birmingham (UAB), 89 pregnant women were enrolled between March 2015 and August 2018. Characteristics of the enrolled study subjects are shown in Table I. Women were enrolled in the first trimester and participated in three study clinic visits during pregnancy (<15 weeks 0 days gestation, 20-22 weeks, 30-32 weeks) and one visit at 4 months post-partum. All asthmatic patients had a prior diagnosis of asthma as outlined in the GINA guidelines (Global Strategy for Asthma Management and Prevention; http://www.ginasthma.org/) (61). Women with asthma were considered “well controlled” (WCAP, n=24) if their initial score on the Asthma Control Test was greater than or equal to 20, while lower scoring women with asthma were considered “poorly controlled” (AP, n=48; WCAP, n=16; total AP/WCAP n=64) and women with no asthma history comprised the “no asthma” group (HP, n=25). The study was approved by UAB review board (IRB-140321010), and written informed consent was obtained from all participants.

### Isolation of peripheral blood mononuclear cells (PBMCs)

Peripheral blood (10 ml) was collected from each participant using BD Vacutainer tubes containing acid-citrate-dextrose anticoagulant, solution A (ACD-A; BD, Franklin Lakes, NJ). 1 ml blood was aliquoted to a 1.5 ml Eppendorf tube and centrifuged at 2000 x g for 15 min. The plasma was collected into a new 1.5 ml tube and stored at −80°C for future experiments. Peripheral blood mononuclear cells (PBMCs) were isolated by density gradient centrifugation with Ficoll-Paque PLUS density gradient media (GE Healthcare, Chicago, IL) from 9 ml of peripheral blood as previously described (Ref: Current Protocols in MicrobiologyA.4C.1-A.4C.9, August 2007). Freshly isolated PBMCs were utilized for flow cytometry analysis.

### Flow Cytometry

PBMCs were blocked in RPMI 1640 (Gibco, Invitrogen, UK) supplemented with 10% human serum followed by staining with relevant antibodies. Anti-Human CD163 (clone: eBioGHI/61 (GHI/61)-PE, anti-Human HLA-DR (clone: LN3)-APC, anti-Human CD15 (clone: HI98)-eFluor® 450, anti-Human CD16 (clone: eBioCB16)– FITC, anti-Human CD11c (clone: 3.9)-PE-Cyanine5, anti-Human CD14 (clone: 61D3)-PE-Cyanine7, anti-Human ILT-2 (clone: 61D3)–PE antibodies were purchased from Life Technologies (Carlsbad, CA). Anti-CD11b (clone ICRF44)-APC-Cy7 was bought from BD Bioscience (San Jose, CA). Anti-Human CD66b (clone: G10F5)-PerCP-Cy5.5 was purchased from Biolegend (San Diego, CA). Anti-Human ILT-4–Alexa Fluor 700 was from R&D Systems (Minneapolis, MN). Data were collected with BD LSR-II flow cytometer (Franklin Lakes, NJ) and analyzed with FlowJo software (Tree Star, Ashland, OR).

### IDO activity assay

The bicinchoninic acid assay (BCA assay) was performed to determine protein concentration in sera (100 dilutions in PBS-pH6.5) of asthmatic, well controlled and healthy pregnant women. Volume of sera was calculated based on 20 μg protein and PBS was added (pH 6.5) to make final volume 250μL. 2X IDO reaction buffer was made in presence or absence of Tryptophan (Trp). Tryptophan supplemented (Trp^+^) 2X IDO reaction buffer was prepared with 800 mM Trp, 40 mM ascorbic acid, 20 mM methylene blue and 200 μg/mL catalase in 1X PBS (pH 6.5). Another 2X IDO reaction buffer (Trp^-^) was prepared as described above but without Trp. Equal volume of 2X IDO reaction buffer was mixed with 250 μL of serum samples separately for Trp^+^ and Trp^-^ groups and incubated for 30 minutes at 37°C. 30% TCA was added in Trp^+^ and Trp^-^ sample tubes and incubated for 30 minutes at 52°C in water bath. The tubes were then centrifuged at 10,000 x g for 5 minutes at 4°C. L-Kynurenine standards were made for 30,000-3 μM range. 3 replicates of 100 μL Supernatant of Trp^+^ and Trp^-^ samples were added in 96 well plates and topped with equal volume of Ehrlich’s reagent to each wells including L-Kyn standards and incubated 10 minutes at RT. IDO activity in nmole/hr was calculated by measuring absorbance at 480 nm.

### Quantitation of s-HLAG by ELISA

Soluble major histocompatibility complex, class I, G (s-HLAG) was measured by commercially available ELISA kit from MYBioSource (San Diego, CA). The assay was performed according to manufacturer’s protocol. Results were shown as a concentration of s-HLAG (U/mL) in the plasma samples.

### Fractional exhaled nitric oxide (FeNO) test

Fractional exhaled nitric oxide is a measure of airway inflammation. In the B-WELL-Mom study, FeNO was measured using the NIOX Vero (Circassia), a quantitative, non-invasive, simple and safe method to measure the levels of FeNO in asthma patients and other conditions characterized by airway inflammation. The NIOX VERO was operated with a 10 second exhalation time. FeNO was collected from women at each of the four study visits and recorded on the study visit forms.

### Statistical analysis

The distributions of MDSCs subgroups (Mo-MDSCs and Gr-MDSCs), IDO activities, sHLA-G levels and FeNO measurements were inspected and summarized by means, 2.5 percentiles and 97.5 percentiles, stratified by visits and patient groups (HP and AP/WCAP). Mixed effects models were applied to examine associations between each outcome and visits, patient groups, and their interaction. Pairwise comparisons were conducted to compare visits within groups and groups within visits. Statistical significance (*p* < 0.05) was determined using the *F* statistic. Pearson correlation coefficients were calculated to measure linear correlations between outcomes. The results were visualized in heat plots for correlation matrices by visits and patient groups. The analyses were implemented using SAS software (version 9.4; SAS Institute Inc., Cary, NC) and R software (version 3.6.1).

## Results

### Dynamics of circulating MDSCs during pregnancy and post-partum

To investigate whether frequencies of circulating MDSC subpopulations change during pregnancy and post-partum, we characterized the percentages of two monocytic subsets, CD14^+^HLA-DR^neg^CD11b^+^CD16^+^CD15^neg^CD66b^neg^ and CD14^+^HLA-DR^neg^CD11b^+^CD16^neg^CD15^neg^CD66b^neg^, classified as Mo-MDSCs and HLA-DR^neg^CD15^+^CD16^+^CD66b^+^CD11b^+^ as Gr-MDSCs in PBMCs isolated from healthy pregnant controls (HP) and asthmatic pregnant subjects including both “poorly controlled” and “well controlled” (AP/WCAP) groups. The gating strategy for identifying these circulating myeloid cell populations is shown in Supplementary Figure-1 and flow cytometry controls are shown in Supplementary Figure −2. In HP subjects, compared to visit 1, the percentage of CD16^+^ Mo-MDSCs in visit 3 decreased marginally (p = 0.05); but showed an increasing trend in visit 4 compared to that of visit 3 (p = 0.07) **(Figure 1A**, **Table II)**. In AP/WCAP subjects, the percentage of these CD16^+^ Mo-MDSCs decreased significantly in visit 3 compared to that of visit 1 (p<0.0001) and visit 2 (p=0.006), and remained significantly reduced at visit 4 compared to that of visit 1 (p=0.007) **(Figure 1A**, **Table II)**. These differences in circulation dynamics was not observed for the CD16^neg^ Mo-MDSC subset in either HP or AP/WCAP subjects during pregnancy **(Figure 1B**, **Table II)**. In HP subjects, the percentage of Gr-MDSCs in visit 4 decreased significantly compared to that of visit 1 (p=0.02), visit 2 (p=0.003) and visit 3 (p=0.0003) **(Figure 1C**, **Table II)**. Interestingly, in AP/WCAP subjects, the percentage of Gr-MDSCs in visit 4 decreased compared to visit 1 (p=0.03) and visit 2 (p=0.008), while decreased only marginally compared to that of visit 3 (p = 0.06) **(Figure 1D**, **Table II).** Comparisons of percentage of CD16^+^ Mo-MDSCs between AP/WCAP and HP subjects showed an increased trend in the asthmatics compared to HP subjects in visit 1 (p = 0.08) and visit 2 (p = 0.06) **(Figure 1E)**; these differences were not observed in visits 3 & 4. Visit differences between study groups were not noted for CD16^neg^ Mo-MDSCs **(Figure 1F)**. Additionally, percentage of Gr-MDSCs between HP and AP/WCAP groups in visit 1, 2 and 3 were not significantly different **(Figure 1G)**. However, during visit 4, the percentage of Gr-MDSCs in AP/WCAP subjects increased compared to that of HP subjects **(Figure 1F).** These results indicate that the dynamics of Mo-MDSCs and Gr-MDSCs in circulation is different across the 4 visits in AP/WCAP compared to HP subjects.

**Figure 1.**
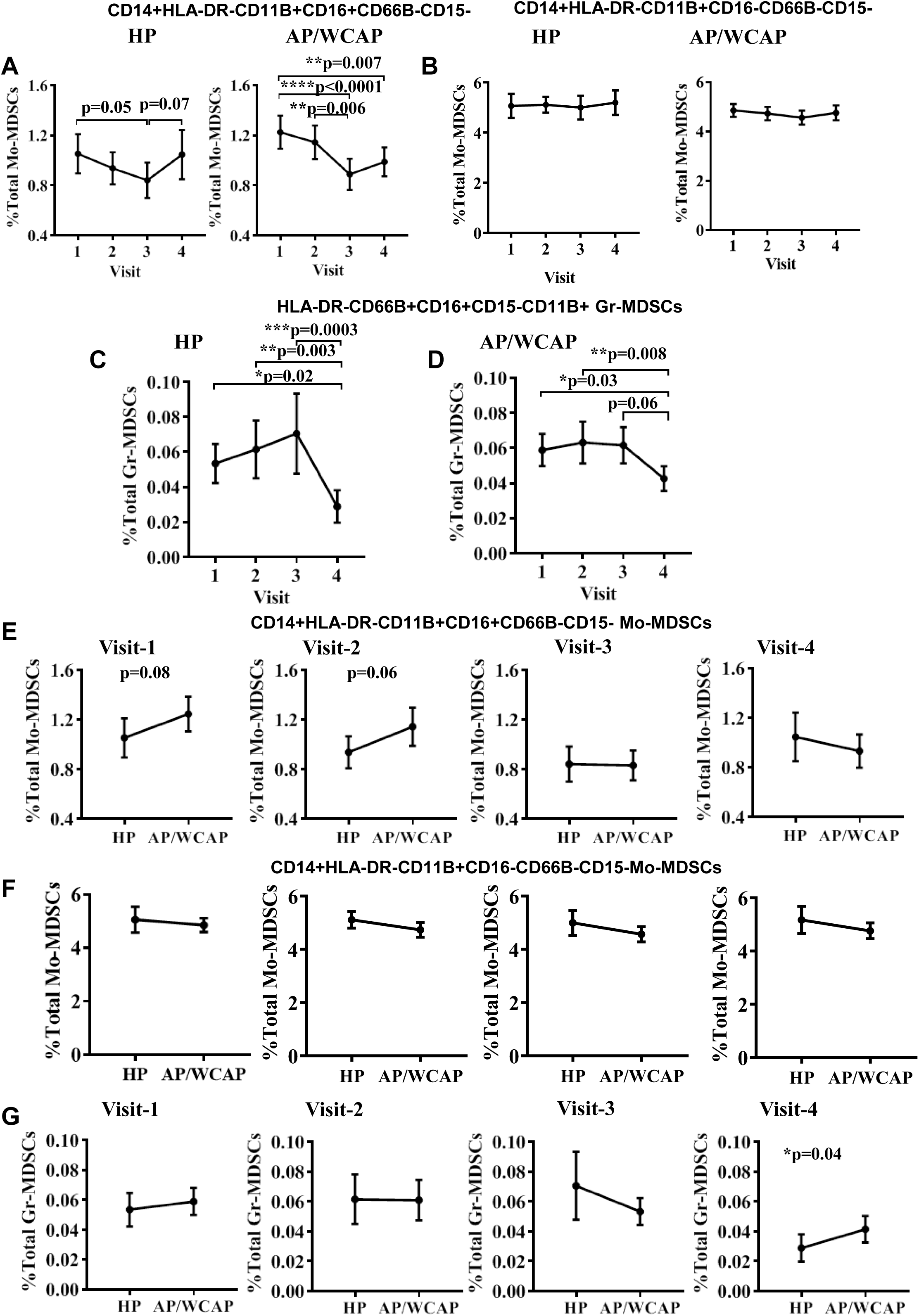
Frequencies of circulating MDSC, IDO activity, sHLA-G levels and eNO measurements among four visits in HP subjects and AP/WCAP patients. PBMCs were isolated by density gradient centrifugation with Ficoll-Paque PLUS density gradient media from about 9 ml of peripheral blood. Freshly isolated PBMCs were utilized for the following flow cytometry analysis. **A.** The percentages of circulating CD14^+^HLA-DR^neg^CD11b^+^CD16^+^CD15^neg^CD66b^neg^ Mo-MDSCs during 4 visits in HP subjects (n=25) and AP/WCAP patients (n=64). **B.** The percentages of circulating CD14^+^HLA-DR^neg^CD11b^+^CD16^neg^CD15^neg^CD66b^neg^ Mo-MDSCs during 4 visits in HP subjects (n=25) and AP/WCAP patients (n=64). **C.** The frequencies of circulating HLA-DR^neg^CD15^+^CD16^+^CD66b^+^CD11b^+^ Gr-MDSCs during 4 visits in HP subjects (n=25) **D.** The frequencies of circulating HLA-DR^neg^CD15^+^CD16^+^ CD66b^+^CD11b^+^ Gr-MDSCs during 4 visits in AP/WCAP patients (n=64). **E.** Comparison of percentages of circulating CD14^+^HLA-DR^neg^CD11b^+^CD16^+^ CD15^neg^CD66b^neg^ Mo-MDSCs during each of the 4 visits in HP subjects (n=25) versus AP/WCAP patients (n=64). **F**. Comparison of percentages of circulating CD14^+^HLA-DR^neg^CD11b^+^CD16^neg^ CD15^neg^CD66b^neg^ Mo-MDSCs during each of the 4 visits in HP subjects (n=25) versus AP/WCAP patients (n=64). **G.** Comparison of percentages of circulating HLA-DR^neg^CD15^+^CD16^+^CD66b^+^CD11b^+^Gr-MDSCs during each of the 4 visits in in HP subjects (n=25) versus AP/WCAP patients (n=64).

### Alteration of IDO activities during pregnancy of HP and AP/WCAP subjects

Indoleamine 2,3-dioxygenase (IDO), a heme-containing enzyme, catalyzes the conversion of the essential amino acid tryptophan (Trp) into the metabolic byproduct kynurenine (Kyn). To investigate whether IDO activity changes during pregnancy, we measured IDO activity in plasma from HP and AP/WCAP study groups. In plasma samples from HP, the IDO activity in visit 2 showed decreased trend compared to visit 1 samples **(Figure 2A)**. However, this difference in IDO activity between visits, was not observed in AP/WCAP group **(Figure 2B).** When IDO activity was compared across study groups, IDO activity in AP/WCAP plasma samples in visit 1 was marginally increased compared to HP samples (p = 0.05) (**Figure 2C**), with no change observed during visits 2, 3 & 4. Together, these results suggest that IDO activity of AP/WCAP patients increased compared to HP subjects only in early stage of pregnancy, while it remains unchanged in later stages of pregnancy **(Figure 2C).**

**Figure 2.**
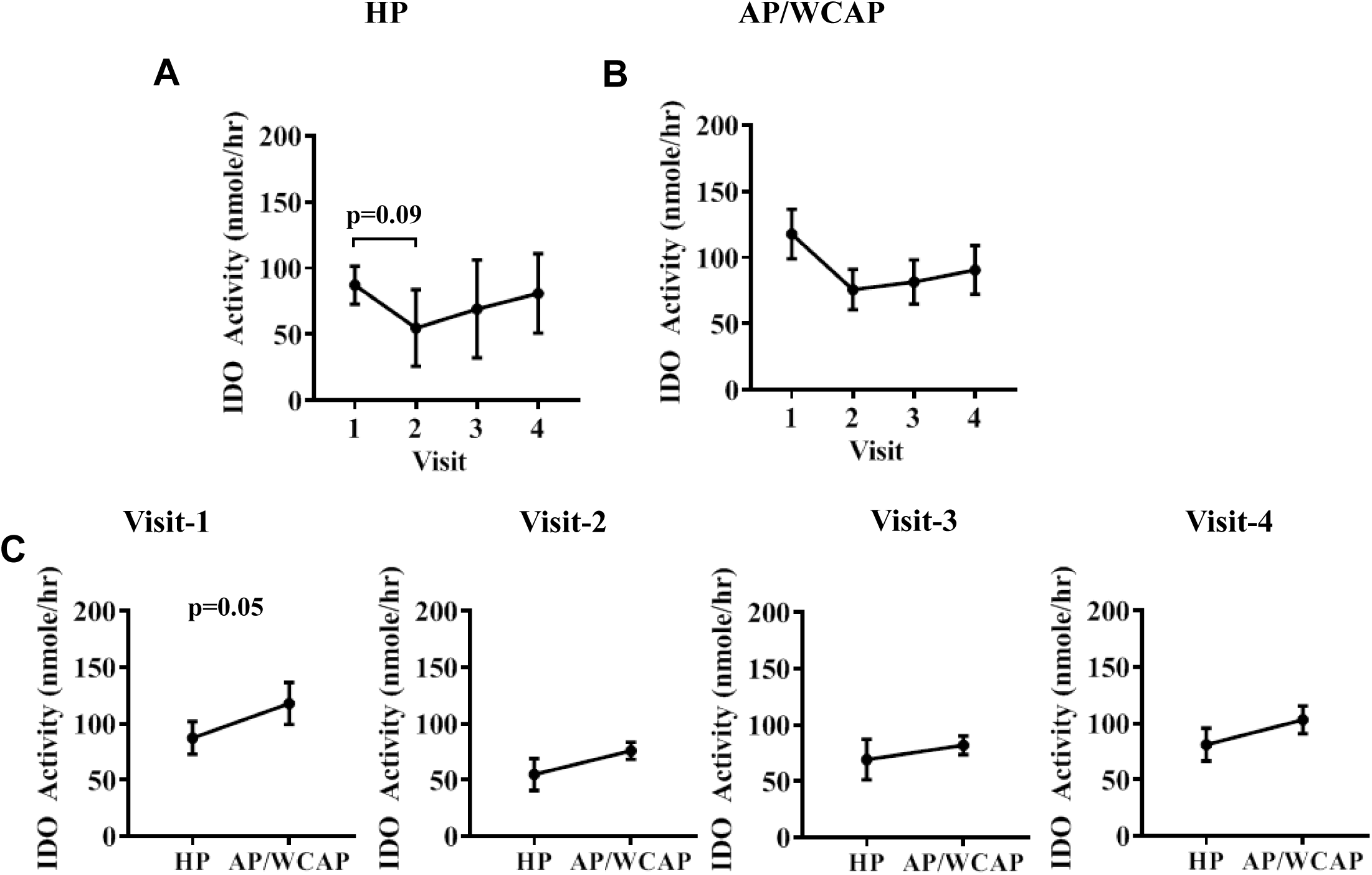
IDO activity during pregnancy comparing HP and AP/WCAP subjects. IDO activity in plasma was quantitated from HP and AP/WCAP groups. **A.** IDO activity in HP subjects (n=25) during 4 visits. **B.** IDO activity in AP/WCAP patients (n=64) during 4 visits. **C.** Comparison of IDO activity in HP subjects (n=25) versus AP/WCAP patients (n=64) during each of the 4 visits.

### Modulation of sHLA-G levels during pregnancy in HP and AP/WCAP subjects

HLA-G molecules, both in membrane-bound and in soluble forms, play a central role in modulation of immune responses during pregnancy and cause accumulation of MDSCs during pregnancy (48,62). To investigate a potential role for sHLA-G in MDSC dynamics, we measured sHLA-G plasma levels in HP and AP/WCAP samples. In both HP and AP/WCAP study groups, the plasma sHLA-G levels were elevated in visit 2 and visit 3 compared to visit 1, whereas the sHLA-G level was reduced in visit 4 as compared to that of visit 2 or visit 3 **(Figure 3A** & **3B).** The sHLA-G level at visit 3 was more variable amongst HP subjects than the AP/WCAP subjects, but no significant difference was identified between HP and AP/WCAP samples in all four visits **(Figure 3C)**. These results indicate that both AP/WCAP and HP subjects have similar pattern of changes in sHLA-G during the 4 study visits across the pregnancy period.

**Figure 3.**
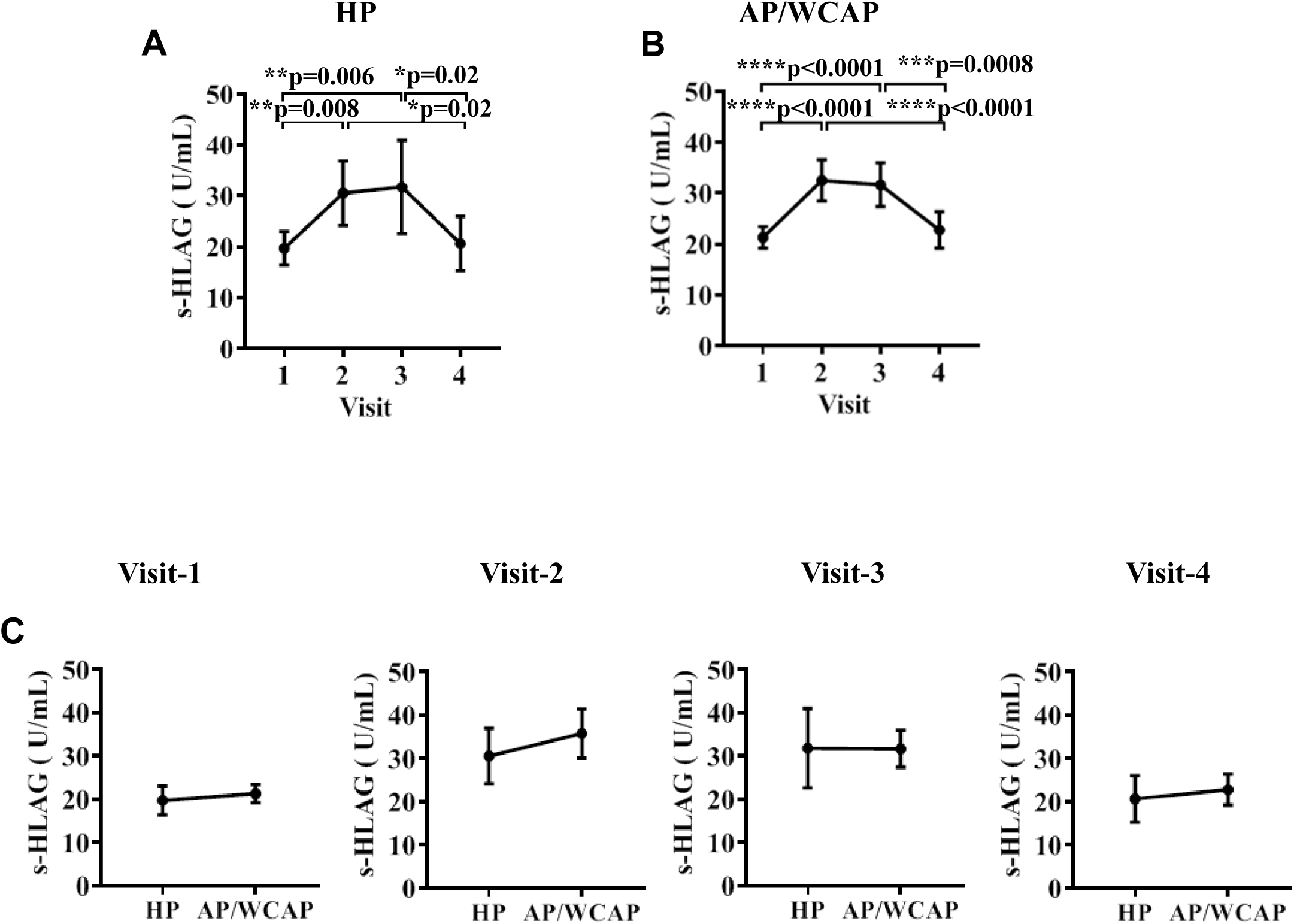
Alterations in sHLA-G levels during pregnancy comparing HP and AP/WCAP subjects. sHLA-G plasma levels were measured by ELISA in HP and AP/WCAP samples. **A.** sHLA-G levels in HP subjects (n=25) during 4 visits. **B.** sHLA-G levels in AP/WCAP patients (n=64) during 4 visits. **C.** Comparison of sHLA-G levels in HP subjects (n=25) versus AP/WCAP patients (n=64) during each of the 4 visits.

### FeNO measurements among four visits in HP subjects and AP/WCAP patients

Fractional exhaled nitric oxide (eNO) measurement is a good surrogate marker for airway inflammation. We evaluated potential FeNO changes during pregnancy in asthmatics compared to control groups, in HP and AP/WCAP samples. In HP subjects, there was no change in FeNO levels during 4 visits **(Figure 4A).** However, in AP/WCAP patients, during visit 1, the FeNO showed an increasing trend (p = 0.08) compared to HP subjects **(Figure 4C)**, but during visit 2, this difference became significant (p=0.03). FeNO level showed a decreased trend in visit 3 compared to that of visit 1 (p = 0.09) **(Figure 4B),** and was only marginally elevated in AP/WCAP patients compared to HP subjects during visit 4 (p = 0.05) **(Figure 4C)**. These results indicate that FeNO level in AP/WCAP patients increased compared to HP subjects in all visits except for visit 3.

**Figure 4.**
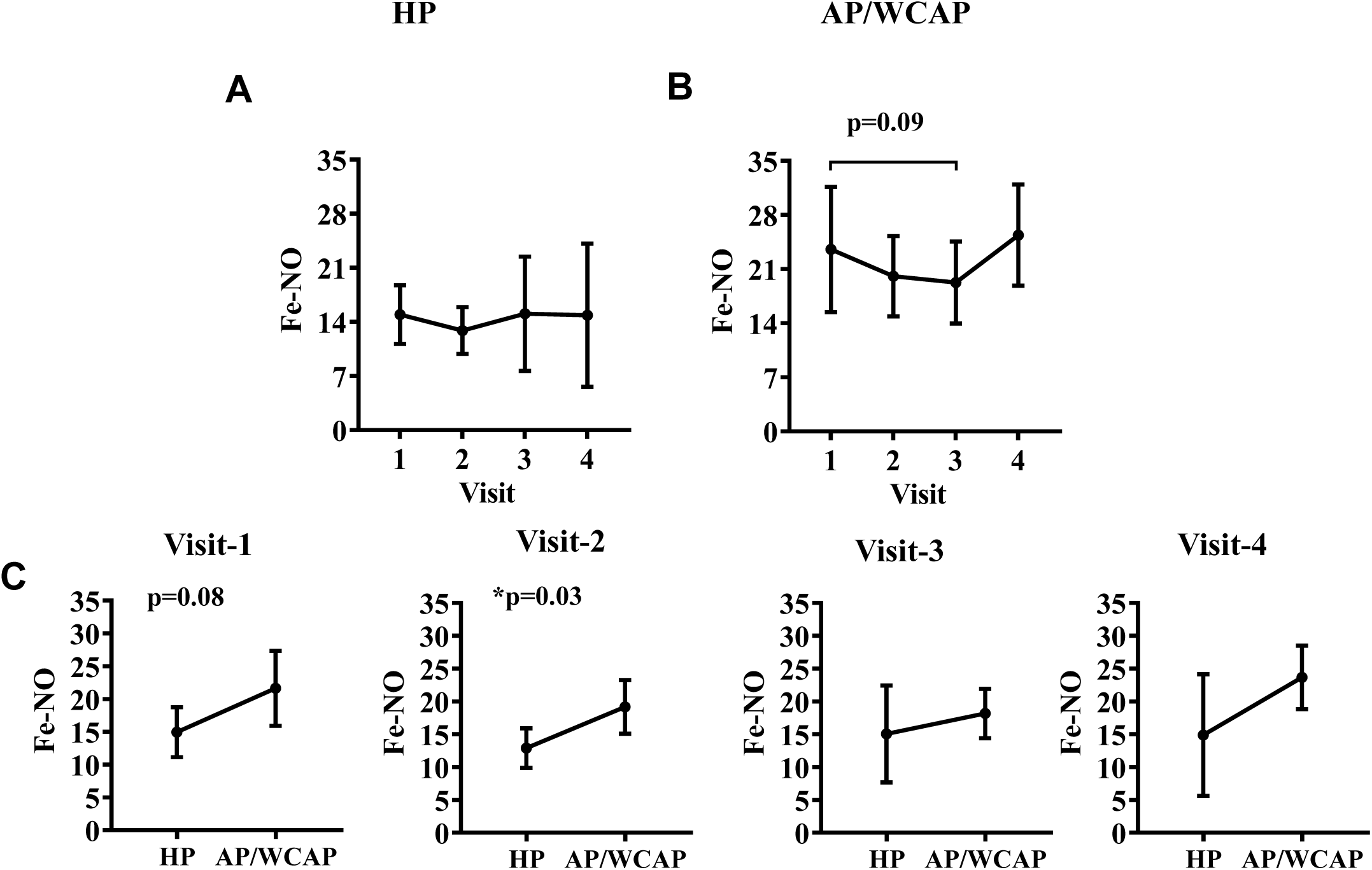
Modulation of FeNO levels during pregnancy comparing HP and AP/WCAP subjects. FeNO levels were measured in HP and AP/WCAP samples. **A.** FeNO levels in HP subjects (n=25) during 4 visits. **B.** FeNO levels in AP/WCAP patients (n=64) during 4 visits. **C.** Comparison of FeNO levels in HP subjects (n=25) versus AP/WCAP patients (n=64) during each of the 4 visits.

### Correlation analyses among frequency of circulation MDSCs, sHLA-G, IDO and FeNO

Positive correlations were observed between circulating CD16^+^ Mo-MDSC subset and Gr-MDSCs in both early and late stage of pregnancy among AP/WCAP patients (Correlation Coefficient: visit 1 = 0.46, visit 3 = 0.35) (**Figure 5A**). In HP subjects, however, positive correlation between these two subsets was only observed in visit 2 (Correlation Coefficient = 0.36) **(Figure 5A)**. Interestingly, in AP/WCAP subjects, the CD16^neg^ Mo-MDSC subset did not correlate with Gr-MDSC during any of the visits; while positive correlation was noted in HP subjects during late stage of pregnancy (visit 2 and visit 3) (**Figure 5B**).

**Figure 5.**
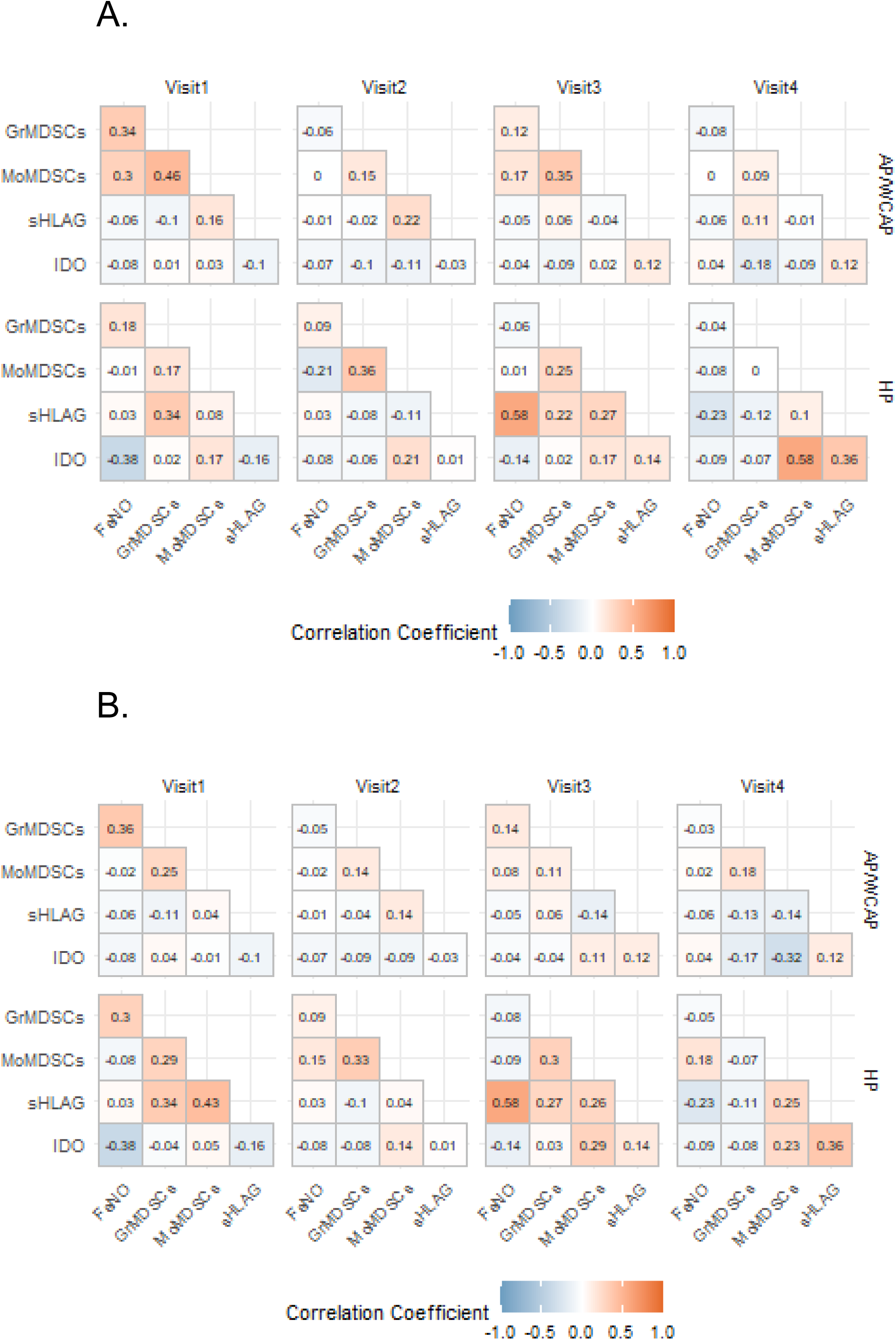
**Correlation analyses of MDSC frequencies with sHLA-G, FeNO, and IDO activity during pregnancy and post-partum in AP/WCAP and HP subjects.** A. Correlation analyses of CD16^+^ Mo-MDSC, Gr-MDSC, FeNO, sHLA-G and IDO activity during each visit of pregnancy. B. Correlation analyses of CD16^neg^ Mo-MDSC, Gr-MDSC, FeNO, sHLA-G and IDO activity during each visit of pregnancy.

We then correlated MDSC frequencies with indicators of pregnancy and asthma. The Gr-MDSC frequencies positively correlated with FeNO in early pregnancy at visit 1 among AP/WCAP subjects (Correlation Coefficient = 0.34), and not in HP subjects **(Figure 5)**. In HP subjects, IDO activity negatively correlated with FeNO (Correlation Coefficient = −0.38) during early pregnancy at visit 1, while positive correlation of sHLA-G level (Correlation Coefficient = 0.58) and frequency of CD16^+^ Mo-MDSCs (Correlation Coefficient = 0.36) with IDO activity levels was observed at 4 months post-partum (visit 4) among only HP group **(Figure 5)** and not in AP/WCAP group. sHLA-G levels positively correlated with FeNO at visit 3 among HP subjects and not in AP/WCAP subjects (Correlation Coefficient = 0.58) **(Figure 5).** Additionally, positively correlation between sHLA-G level and Gr-MDSCs (Correlation Coefficient = 0.34) was identified in early pregnancy at visit 1 of HP subjects and not AP/WCAP subjects **(Figure 5)**. Interestingly, in HP subjects, the frequency of CD16^neg^ Mo-MDSCs correlated positively with sHLA-G levels in early pregnancy (Correlation Coefficient = 0.43), but did not correlate at 4 months post-partum with IDO activity as observed with the CD16^+^ Mo-MDSC. The differences in correlation signatures amongst frequencies of Gr-MDSCs, Mo-MDSCs, and indicators of pregnancy and asthma such as sHLA-G, IDO activity and FeNO between the two study groups, suggests their differential functional relationships comparing AP/WCAP and HP subjects.

We then determined Visit effects and Group effects of these measures to evaluate potential interactions **(Figure 6).** As shown in **Figure 6**, the Visit effects were significantly different for sHLA-G (p<0.0001), Mo-MDSCs (p = 0.006) and Gr-MDSCs (p = 0.0004), however, the Group effect was significant for FeNO (p = 0.0004); significant interaction between these measures were not identified.

**Figure 6.**
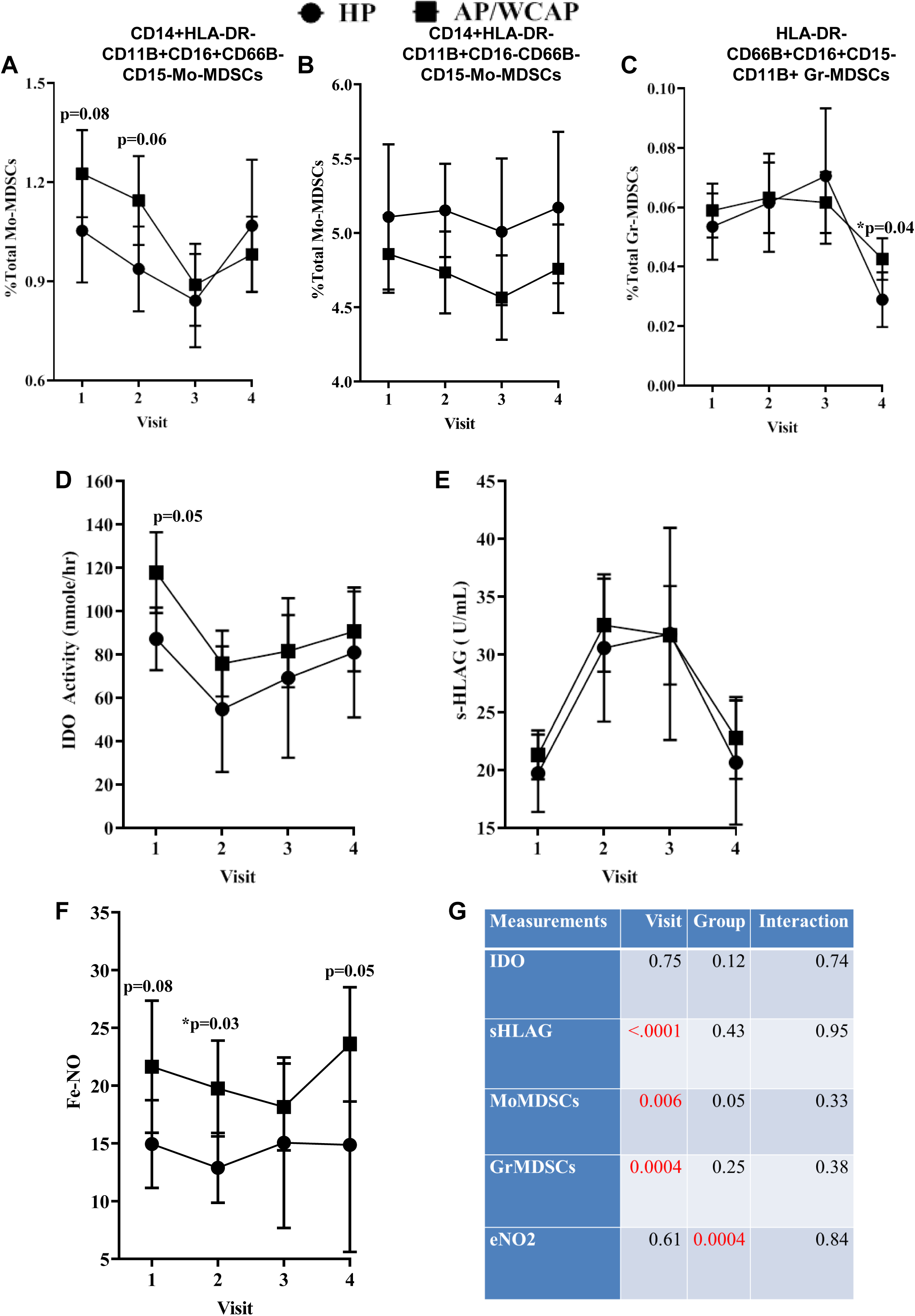
**Comparison of Dynamics of circulating MDSC frequencies and indicators of pregnancy in HP and AP/WCAP subjects.** (A) Dynamics of CD16^+^ Mo-MDSCs during each of the 4 visits of pregnancy in HP and AP/WCAP subjects. (B) Dynamics of CD16^neg^ Mo-MDSCs during each of the 4 visits of pregnancy in HP and AP/WCAP subjects. (C) Dynamics of Gr-MDSCs during each of the 4 visits of pregnancy in HP and AP/WCAP subjects. (D) Changes in IDO activity during each of the 4 visits of pregnancy in HP and AP/WCAP subjects. (E) Changes in sHLA-G levels in plasma during each of the 4 visits of pregnancy in HP and AP/WCAP subjects. (F) Changes in FeNO levels in plasma during each of the 4 visits of pregnancy in HP and AP/WCAP subjects. (G) Table showing Interaction of parameters in A-F.

## Discussion

In this study, we report that there are differences in dynamics of circulating MDSCs in asthmatics and healthy controls during various stages of pregnancy. MDSC dynamics correlate differentially with pregnancy indicators in HP versus AP/WCAP. Our observations that the percentage of circulating Gr-MDSCs increases over pregnancy during early gestation and declines during the last stage of pregnancy in HP subjects is consistent with earlier reports in pregnant mice and in humans as Gr-MDSCs have been shown to decline post-partum (48,62,63). Although the decline of Gr-MDSCs at Visit 4 was also significant in AP/WCAP just as in HP subjects, the increase in their frequency in circulation during early gestation was not as robust as in HP subjects, with AP/WCAP subjects having both higher baseline levels and higher frequencies at Visit 2. As Gr-MDSC increases have been implicated in fetal immune tolerance and maintenance of pregnancy in HP subjects, this may be affected in AP/WCAP as the MDSC frequencies in circulation during early gestation are not as robust as in HP subjects. The increase in baseline frequencies of Gr-MDSCs in AP/WCAP also may contribute to this lack of significant change and potentially be an indicator of underlying oxidative stress/inflammation during early pregnancy. The decline in frequencies of CD14^+^HLA-DR^neg^CD11b^+^CD16^+^ Mo-MDSC subset was similar in both HP and AP/WCAP groups from visit 1 through visit 3, but these frequencies returned to baseline levels in HP at post-partum, whereas in AP/WCAP, it remained significantly reduced from visit 1 levels. Similar to that of Gr-MDSCs, this change in dynamics may indicate underlying oxidative stress/inflammation in AP/WCAP subjects. The FeNO levels are higher at baseline and at visit 2 in AP/WCAP compared to HP. This is consistent with higher FeNO levels commonly seen in asthmatics. The higher FeNO levels at visits 1 & 2 in AP/WCAP correlate with higher levels of Gr-MDSCs and Mo-MDSCs and in AP/WCAP. Whereas, MDSC subsets have been shown to produce and utilize NO for immune regulation, it remains to be determined if these myeloid-lineage ells directly contribute to FeNO. Although airway MDSC subsets have been enumerated and their function evaluated in the airways of asthmatics (17), they have not been assessed in pregnancy. The dynamics of CD14^+^HLA-DR^neg^CD11b^+^CD16^+^ Mo-MDSCs is reciprocal to that of Gr-MDSCs at visit 3. It is unclear if circulatory pools of different subpopulations with different function exist, or whether the local microenvironment facilitates their transdifferentiation, which remains to be determined.

The observed positive correlations between sHLA-G and accumulation of Gr-MDSCs, both at visits 1 and 3, as well as moderate correlation of sHLA-G with Mo-MDSCs at Visit 3 only in HP subjects are also consistent with sHLA-G’s known role in maternal-fetal tolerance. Lack of this positive correlation in AP/WCAP may influence maternal-fetal tolerance and potential complications during pregnancy in asthmatics. Additionally, sHLA-G significantly correlated with FeNO only in HP subjects in Visit 3, which was significantly different from AP/WCAP subjects. In HP subjects, during post-partum visit (Visit 4), activity of IDO, a major player in maternal-fetal interface, positively correlated with sHLA-G and Mo-MDSCs. This positive correlation was absent in AP/WCAP subjects suggesting again the maternal-fetal tolerance may be affected in AP/WCAP subjects. Our studies thus highlight a potential functional relationship between circulating MDSC dynamics and indicators of maternal-fetal tolerance during human pregnancy and suggest the potential utility of MDSC subsets to serve as biomarkers of maternal-fetal tolerance in asthmatic pregnant women.

## Data Availability

All data produced in the present study are available upon reasonable request to the authors.

## ACKNOWLEDGEMENTS

The authors would like to acknowledge the UAB Comprehensive Flow Cytometry Core Facility (NIH P30 AI27667, NIH CA013148), for their help in data collection.

## FUNDING STATEMENT

Research reported in this publication was supported by HHSN275201300014C (awarded to A.S. and J.R.B) and HL128502 awarded to JSD.

---

**Table I.**
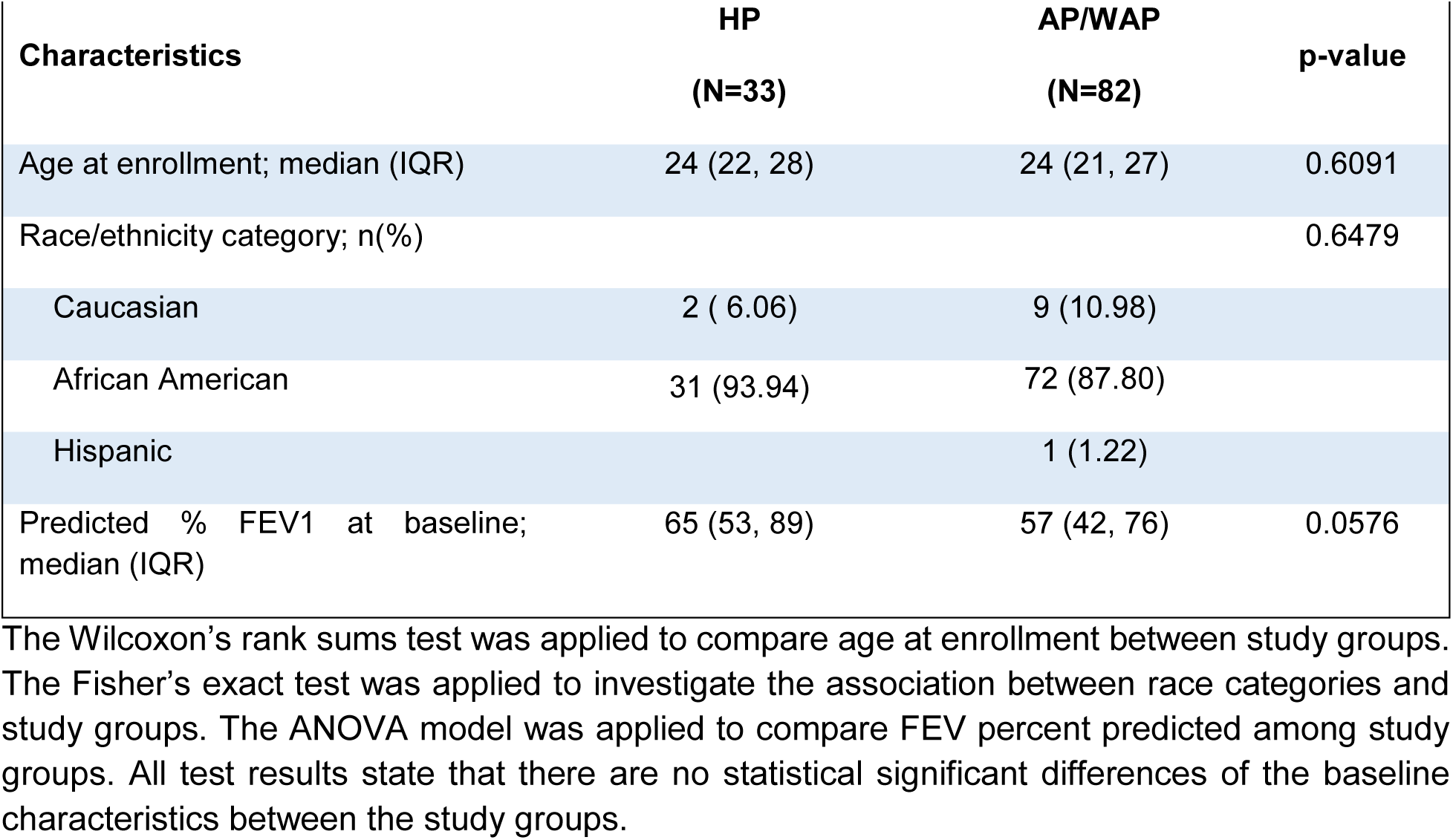
Characteristics of Enrolled Study Subjects

| **Table I: Characteristics of Enrolled Study Subjects** |  |  |  |
| --- | --- | --- | --- |
| **Characteristics** | **HP**<br>**(N=33)** | **AP/WAP**<br>**(N=82)** | **p-value** |
| Age at enrollment; median (IQR) | 24 (22, 28) | 24 (21, 27) | 0.6091 |
| Race/ethnicity category; n(%) |  |  | 0.6479 |
| Caucasian | 2 ( 6.06) | 9 (10.98) |  |
| African American | 31 (93.94) | 72 (87.80) |  |
| Hispanic |  | 1 (1.22) |  |
| Predicted % FEV1 at baseline;<br>median (IQR) | 65 (53, 89) | 57 (42, 76) | 0.0576 |

The Wilcoxon's rank sums test was applied to compare age at enrollment between study groups. The Fisher's exact test was applied to investigate the association between race categories and study groups. The ANOVA model was applied to compare FEV percent predicted among study groups. All test results state that there are no statistical significant differences of the baseline characteristics between the study groups.

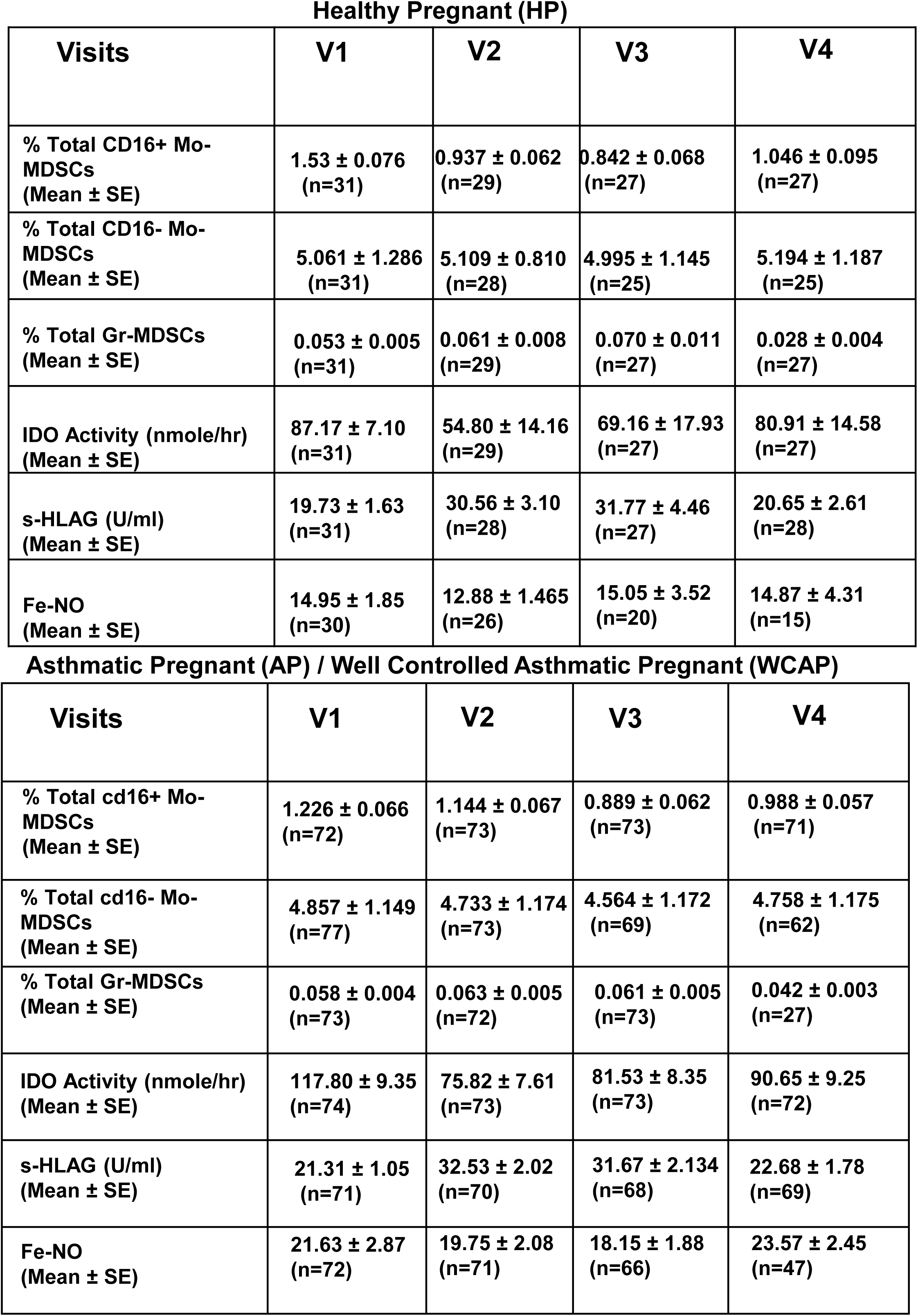

Healthy Pregnant (HP)
| Visits | V1 | V2 | V3 | V4 |
| --- | --- | --- | --- | --- |
| % Total CD16+ Mo-MDSCs<br>(Mean ± SE) | 1.53 ± 0.076<br>(n=31) | 0.937 ± 0.062<br>(n=29) | 0.842 ± 0.068<br>(n=27) | 1.046 ± 0.095<br>(n=27) |
| % Total CD16- Mo-MDSCs<br>(Mean ± SE) | 5.061 ± 1.286<br>(n=31) | 5.109 ± 0.810<br>(n=28) | 4.995 ± 1.145<br>(n=25) | 5.194 ± 1.187<br>(n=25) |
| % Total Gr-MDSCs<br>(Mean ± SE) | 0.053 ± 0.005<br>(n=31) | 0.061 ± 0.008<br>(n=29) | 0.070 ± 0.011<br>(n=27) | 0.028 ± 0.004<br>(n=27) |
| IDO Activity (nmole/hr)<br>(Mean ± SE) | 87.17 ± 7.10<br>(n=31) | 54.80 ± 14.16<br>(n=29) | 69.16 ± 17.93<br>(n=27) | 80.91 ± 14.58<br>(n=27) |
| s-HLAG (U/ml)<br>(Mean ± SE) | 19.73 ± 1.63<br>(n=31) | 30.56 ± 3.10<br>(n=28) | 31.77 ± 4.46<br>(n=27) | 20.65 ± 2.61<br>(n=28) |
| Fe-NO<br>(Mean ± SE) | 14.95 ± 1.85<br>(n=30) | 12.88 ± 1.465<br>(n=26) | 15.05 ± 3.52<br>(n=20) | 14.87 ± 4.31<br>(n=15) |

Asthmatic Pregnant (AP) / Well Controlled Asthmatic Pregnant (WCAP)
| Visits | V1 | V2 | V3 | V4 |
| --- | --- | --- | --- | --- |
| % Total cd16+ Mo-MDSCs<br>(Mean ± SE) | 1.226 ± 0.066<br>(n=72) | 1.144 ± 0.067<br>(n=73) | 0.889 ± 0.062<br>(n=73) | 0.988 ± 0.057<br>(n=71) |
| % Total cd16- Mo-MDSCs<br>(Mean ± SE) | 4.857 ± 1.149<br>(n=77) | 4.733 ± 1.174<br>(n=73) | 4.564 ± 1.172<br>(n=69) | 4.758 ± 1.175<br>(n=62) |
| % Total Gr-MDSCs<br>(Mean ± SE) | 0.058 ± 0.004<br>(n=73) | 0.063 ± 0.005<br>(n=72) | 0.061 ± 0.005<br>(n=73) | 0.042 ± 0.003<br>(n=27) |
| IDO Activity (nmole/hr)<br>(Mean ± SE) | 117.80 ± 9.35<br>(n=74) | 75.82 ± 7.61<br>(n=73) | 81.53 ± 8.35<br>(n=73) | 90.65 ± 9.25<br>(n=72) |
| s-HLAG (U/ml)<br>(Mean ± SE) | 21.31 ± 1.05<br>(n=71) | 32.53 ± 2.02<br>(n=70) | 31.67 ± 2.134<br>(n=68) | 22.68 ± 1.78<br>(n=69) |
| Fe-NO<br>(Mean ± SE) | 21.63 ± 2.87<br>(n=72) | 19.75 ± 2.08<br>(n=71) | 18.15 ± 1.88<br>(n=66) | 23.57 ± 2.45<br>(n=47) |

## Notes

### Competing Interest Statement

The authors have declared no competing interest.

### Author Declarations

The study was approved by University of Alabama at Birmingham Institutional Review Board (IRB-140321010) and written informed consent was obtained from all participants

