## Supplementary figures and images for "Regulatory Myeloid-Cell Dynamics In Asthmatics During Pregnancy"

### Supplementary Figures

##### Supplementary Figure 1

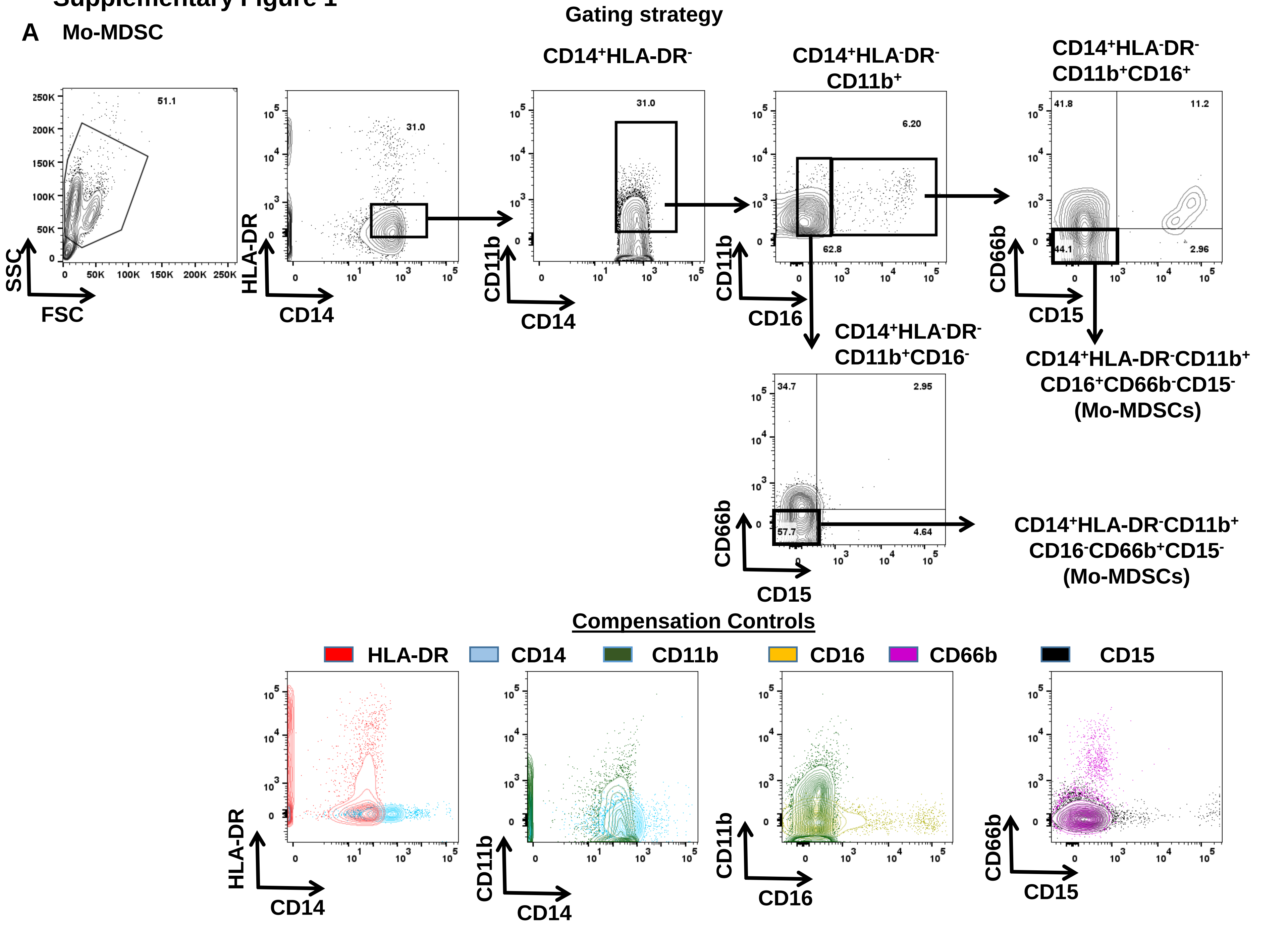

#### Supplementary Figure 2

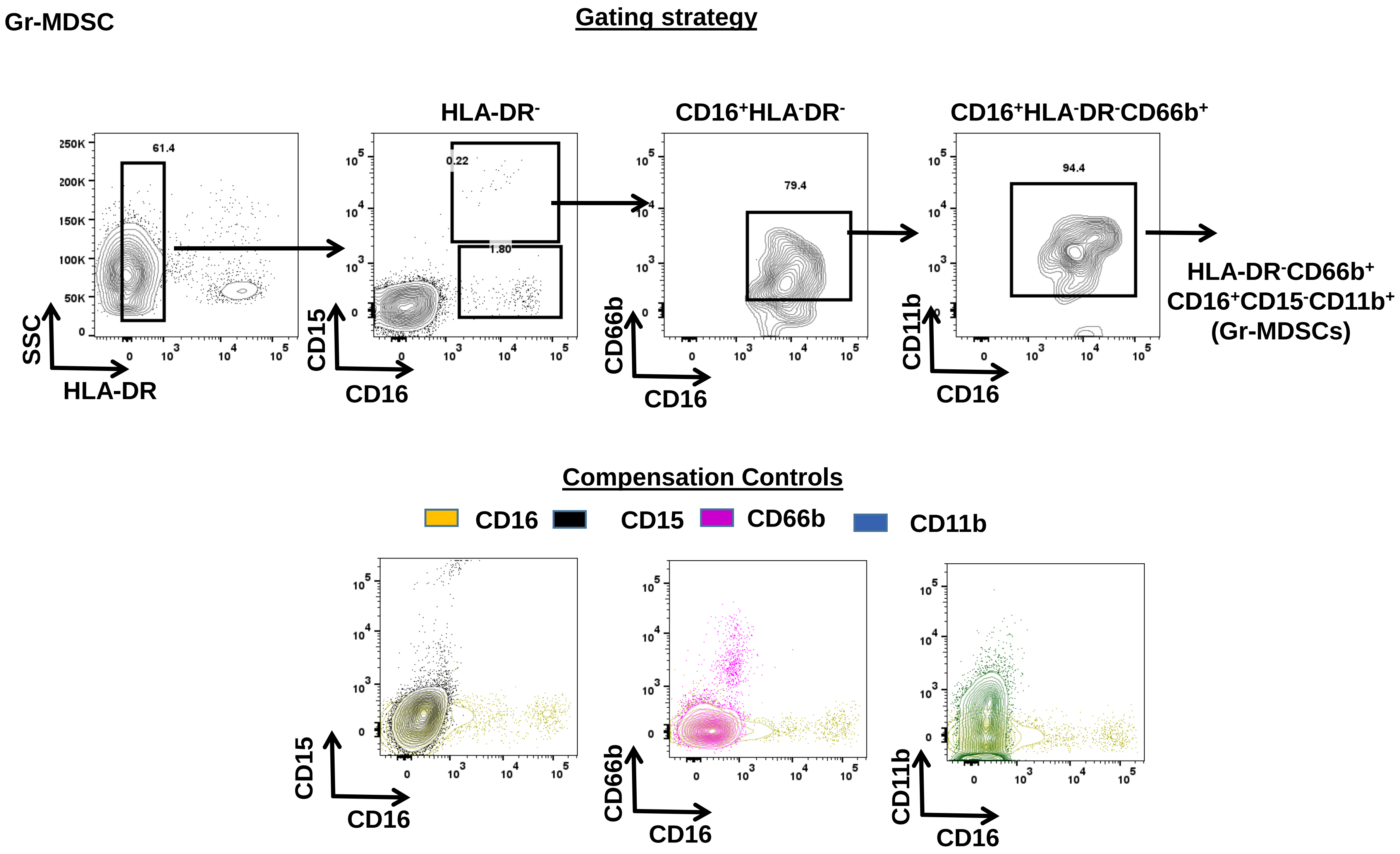
